# No evidence of increased gaming-related problems with long-term use of a video game therapeutic: Exploratory endpoint findings from a randomized controlled trial

**DOI:** 10.64898/2026.03.04.26347656

**Authors:** Lauri Lukka, Joonas J. Juvonen, Satu Palva, Erkki Isometsä, J. Matias Palva

## Abstract

Digital therapeutics for mental health often face low patient engagement, which limits their clinical impact. Interventions that deliver treatment using a video game medium may improve engagement and therapeutic efficacy, but the potential emergence of gaming-related problems remains a safety question and a concern among clinical stakeholders. We examined whether long-term adjunctive use of the video game therapeutic Meliora was associated with changes in gaming-related problems in a three-arm randomized controlled trial. Participants were adults with interview-confirmed major depressive disorder without significant gaming or gambling problems at baseline. The intention-to-treat cohort (*n* = 1,001) had a mean age of 33.4 years (SD 9.3), and 64% were female. From baseline to post-intervention (week 12), Game Addiction Scale (GAS-7) scores decreased in the Meliora group (mean change, −0.63 points; 95% CI, −0.91 to −0.34; *dz* = −0.24; Holm-adjusted *p* < 0.001), whereas no change was observed in the Sham group, which used a highly similar video game therapeutic (mean change, −0.02 points; 95% CI, −0.25 to 0.28; *dz* = −0.01; Holm-adjusted *p* = 0.900). Between-group analyses showed more negative GAS-7 change in the Meliora group than in the Sham group (adjusted mean difference, −0.19 points; 95% CI, −0.30 to −0.08; *d* = −0.13; Holm-adjusted *p* = 0.001), indicating a relatively greater reduction with Meliora, whereas changes were similar between the Meliora and TAU groups (adjusted mean difference = 0.04 points; 95% CI, −0.08 to 0.15; *d* = 0.03; Holm-adjusted *p* = 0.509). Intervention usage time, experienced immersion, and their interaction were not significantly associated with changes in GAS-7 scores in either the Meliora or Sham group. Using a ≥25% deterioration threshold, deterioration occurred in 10.4% of participants in the Meliora group, 15.3% in the Sham group, and 9.8% in the TAU group, with no statistically significant between-group differences. Complementary per-protocol analyses among participants with greater intervention exposure were consistent with the ITT findings. These findings provide no evidence that long-term use of a video game therapeutic was associated with increased gaming-related problems in appropriately screened patients using interventions with safeguards against excessive use.

## Introduction

Digital mental health interventions (DMHIs) can expand treatment reach for mental disorders with effectiveness comparable to that of face-to-face psychotherapies (1,2). Based on DMHIs, several commercial digital therapeutics (DTx) have been developed (3,4). Yet, engagement has remained a major barrier: most patients use these products less than recommended, and completion rates in routine care settings are, on average, only 25% (5). Given that long-term engagement predicts treatment outcomes (5–7), these findings suggest that more compelling digital interventions are required.

As a novel form of DTx, video game therapeutics use a video game medium to deliver therapeutic mechanisms of action (8–10). Video games are highly popular and appealing, with 61% of the U.S. population aged 5–90 playing video games at least one hour a week (11). Compared to conventional digital therapeutics, video game therapeutics can have greatly improved appeal and engagement. This can promote experienced immersion and long-term use, which are prerequisites for intervention efficacy (8,10). However, the video game medium also raises concerns about gaming-related problems as adverse effects for these interventions (12). To the best of our knowledge, no prior randomized trial has tested whether a video game therapeutic intended for long-term use (defined, *e*.*g*., by the EU Medical Device Regulation as continuous use for more than 30 days (13)), increases gaming-related problems.

Here, we asked whether the long-term use of a video game therapeutic influences gaming-related problems as measured by the Game Addiction Scale (GAS-7) (14). This analysis addressed a prespecified exploratory endpoint in a randomized controlled superiority trial of a video game therapeutic for adult major depressive disorder (MDD), which compared an adjunctive video game therapeutic (“*Meliora*”), a highly similar adjunctive comparator device (“Sham”), and treatment-as-usual (TAU) alone (10). In the present study, we evaluated whether the GAS-7 scores changed within and between the three study arms from baseline to post-intervention. In addition, we examined whether intervention usage time, experienced immersion, and background covariates were associated with GAS-7 changes, and evaluated GAS-7 score deterioration.

## Methods

This study investigated whether long-term use of a video game therapeutic for adult MDD was associated with increased symptoms of gaming-related problems. This prespecified exploratory endpoint was evaluated within a randomized, double-blinded, comparator-controlled trial in Finland (ClinicalTrials.gov NCT05426265, June 16, 2022) described elsewhere (10). The trial was approved by the Helsinki University Hospital (HUS) Regional Committee on Medical Research Ethics (HUS/3042/2021) and the Finnish Medicines Agency (FIMEA/2022/002976). All patients provided informed consent digitally.

### Patients

A total of 1,001 patients with MDD were randomized into Meliora (*n* = 337), Sham (*n* = 347), or treatment-as-usual (TAU, *n* = 317) arms. Key inclusion criteria were age 18–65 years, meeting diagnostic criteria for MDD based on a remote structured interview using the Mini International Neuropsychiatric Interview (MINI) version 6.0.0 module A (15). To ensure patient safety, patients with significant problems with or addictions to gambling or gaming were excluded from the study. If the patient was not excluded by other criteria and indicated potential gambling- or gaming-related addiction, gambling-related addiction was assessed using the Problem Gambling Severity Index (PGSI), with exclusion defined as a total score of ≥ 8 (16), while gaming-related problems were assessed with the Game Addiction Scale (GAS-7) so that exclusion was defined as endorsement of four or more items at a frequency of “sometimes” or higher (14). Full inclusion and exclusion criteria, recruitment procedures, and intervention design are detailed elsewhere (10). Throughout the study, the participants could contact the clinical study coordinators via email and phone for support and concerns (17).

### Interventions

The active investigational device, *Meliora*, delivered adaptive executive functions training (EFT) in a single-player video game format without social interaction. The device was explicitly positioned as a video game in the patient-facing materials. The highly similar Sham comparator device was a derivative of Meliora and matched its audiovisual and narrative features but had a reduced EFT training load. Patients in the TAU group were given access to either the Meliora or Sham device in a 1:1 ratio from weeks 12 to 24, during which time patients in the Meliora and Sham groups no longer had access to their assigned interventions. Patients used the interventions at home on their personal computers as an adjunct to TAU, with a recommended total usage time of 48 hours over the 12-week intervention period. The devices were thus intended for long-term use of more than 30 days under the EU Medical Device Regulation (13). The devices included a safeguard to mitigate potential excessive use: intervention use was limited to a maximum of 90 minutes per day.

The per-protocol cohort included participants with at least one post-baseline assessment at week 8 and/or week 12. For Meliora and Sham groups, participants were additionally required to have accumulated at least 24 hours of intervention use during the 12-week intervention period; the intervention-use criterion did not apply to the TAU group, which did not use either intervention between weeks 0 and 12.

### Measurement of gaming-related problems and immersion

Gaming-related problems were measured with the Finnish translation of the 7-item Game Addiction Scale (GAS-7) (14): participants were asked to evaluate their experienced addiction to digital games generally with 7 items scored from 1 (“never”) to 5 (“very often”). The GAS-7 instructions asked participants to consider their digital gaming. The questionnaire instructions did not specifically direct the patient to include or to exclude the assigned intervention; however, the study interventions were explicitly presented to participants as video games during the enrolment process. Item scores were summed to yield a total score ranging from 7 to 35, with higher scores indicating greater gaming-related problems. While the Finnish translation has not been independently validated, a previous systematic review found GAS-7 to be the most widely administered tool for screening and assessment of gaming disorder in both adolescents and adults (18). GAS-7 was administered at baseline (week 0), weeks 4 and 8, post-intervention (week 12), and at follow-up (week 24). Internal consistency of the Finnish GAS-7 was assessed separately at each assessment using Cronbach’s α. As an exploratory assessment of longitudinal psychometric stability, one-factor loadings from standardized item scores and corrected item-total correlations were estimated separately at baseline and week 12 and compared across assessments. Immersion was measured with the Finnish translation of the Immersive Experiences Questionnaire (IEQ) (19), administered at weeks 4, 8, and 12.

### Statistical analyses

The exploratory safety hypothesis prespecified in the clinical investigation plan was that the Meliora and Sham interventions would not be associated with an increase in GAS-7 scores from baseline to week 12. The statistical analysis plan did not specify a dedicated inferential procedure for this exploratory hypothesis. We therefore evaluated the prespecified baseline-to-week-12 contrast separately within the Meliora and Sham groups using two-sided paired-samples t tests. A supportive analysis evaluated the corresponding change in the TAU group. Holm correction was applied across the three within-group tests. To characterize the sensitivity of the intervention-group analyses to potential increases in gaming-related problems, we additionally estimated the minimum mean change detectable with 80% power at a two-sided α of 0.05. Standardized within-group effects were calculated as Cohen’s *d*_*z*_ by dividing the mean within-participant change by the standard deviation of the change scores.

In post hoc supportive analyses, GAS-7 change was compared between the three groups using the same robust linear mixed-effects modelling framework as in the primary and secondary analyses of the main trial (10). Models included change-from-baseline scores at weeks 4, 8, and 12, with fixed effects for time and study arm, adjustment for baseline GAS-7, baseline PHQ-9, age, gender, education, income, and life status, and participant-level random intercepts. Between-group contrasts were evaluated using two-sided tests with Holm correction for multiple comparisons. This differed from the one-sided superiority tests used for efficacy outcomes in the main trial, as no strong a priori hypothesis regarding the direction of between-group differences in GAS-7 change was specified. Residual-standardized effect sizes (*d*) were calculated by dividing the model coefficients by the residual standard deviation.

Other post hoc exploratory analyses examined associations between GAS-7 change from baseline to week 12 and total intervention use, experienced immersion, and their interaction using robust linear regression adjusted for baseline GAS-7, age, gender, education, income, and life status; change from week 12 to week 24; and covariate associations with GAS-7 change. Intervention usage time and immersion scores were standardized within each intervention arm before model fitting; accordingly, their regression coefficients represent effects associated with a 1-SD difference in the respective predictor. Residual-standardized effect sizes for the robust linear regression analyses were calculated by dividing model coefficients by the residual standard deviation. Benjamini–Hochberg false discovery rate correction was applied separately to the within-group follow-up and covariate analyses.

Individual-level deterioration from baseline to week 12 was examined post hoc using relative increases in GAS-7 of ≥25%, and with ≥50% examined as a more stringent sensitivity threshold. Deterioration rates were compared between study arms using pairwise two-sided Fisher exact tests with Holm correction. Participants meeting the ≥25% deterioration criterion were further characterized by comparing baseline GAS-7, baseline PHQ-9, age, and, in the intervention arms, total intervention use and experienced immersion with participants not meeting the criterion using Welch’s two-sample t tests. Benjamini–Hochberg false discovery rate correction was applied jointly across these characterization comparisons; GAS-7 change itself was excluded from this multiplicity correction because it formed the basis of the deterioration classification. Gender was examined separately using logistic regression comparing male and female participants within each study arm, with Benjamini–Hochberg false discovery rate correction across the three arm-specific comparisons. Participants reporting genders other than male or female were excluded from these inferential gender comparisons because of sparse deterioration counts. Logistic regression was additionally used to examine whether standardized intervention use was associated with ≥25% deterioration separately in the Meliora and Sham groups.

Although the per-protocol (PP) population was the prespecified analysis population for most outcomes in the main trial, the present safety analyses were conducted primarily in the full intention-to-treat (ITT) cohort to provide the broadest assessment of potential gaming-related problems among randomized participants. Complementary PP analyses included participants who completed the week 8 or week 12 assessment and, for the Meliora and Sham groups, accumulated at least 24 hours of intervention use during the intervention period.

Finally, in a post hoc analysis addressing whether changes in gaming-related problems tracked changes in depressive symptoms, Pearson correlations between PHQ-9 and GAS-7 change from baseline to week 12 were examined separately by study arm and across the pooled study population.

## Results

All enrolled 1,001 participants were included in the intention-to-treat (ITT) cohort (Meliora: *n* = 337; Sham: *n* = 347; TAU: *n* = 317). The complementary per-protocol (PP) cohort included participants who completed the week 8 or week 12 assessment and, for the Meliora and Sham groups, accumulated at least 24 h of intervention use (Meliora: *n* = 99; Sham: *n* = 96; TAU: *n* = 288). Unless otherwise specified, all analyses reported below were conducted in the ITT cohort. The mean age was 33.4 years (SD 9.3), and 64% were female. Baseline GAS-7 scores were similar between groups (Meliora: mean 10.56, 95% CI, 10.22 to 10.91; Sham: mean 10.38, 95% CI, 10.04 to 10.73; TAU: mean 10.50, 95% CI, 10.15 to 10.87). As reported earlier (10), we observed no differences between Meliora and Sham groups in terms of average total intervention use time (18.4 h, SD 21.6; 18.2 h, SD 21.2, respectively, *p* = 0.94), or experienced immersion (118.9, SD 27.1; 115.9, SD 27.0; *p* = 0.25).

### Analysis of the prespecified change-from-baseline hypothesis

The Finnish translation of the GAS-7 showed acceptable internal consistency across assessments, with Cronbach’s α ranging from 0.723 to 0.761 (baseline, *α* = 0.734; week 4, *α* = 0.723; week 8, *α* = 0.761; week 12, *α* = 0.751; week 24, *α* = 0.734). Factor loading profiles were highly similar between baseline and week 12 (*r* = 0.981; mean absolute loading difference = 0.028; maximum = 0.071), as were item-total correlation profiles (*r* = 0.968), with no indication of substantial changes in the scale’s psychometric properties over this period.

From baseline to post-intervention (week 12), mean GAS-7 scores did not increase in either intervention group (Figure 1). GAS-7 scores decreased by 0.63 points in the Meliora group (95% CI, −0.91 to −0.34; *d*_*z*_ = −0.24; Holm-adjusted *p* = 4.2×10^−5^), whereas no change was observed in the Sham group (mean change, −0.02 points; 95% CI, −0.25 to 0.28; *d*_*z*_ = −0.01; Holm-adjusted *p* = 0.90). A post hoc sensitivity analysis indicated that the paired change analyses had 80% power at a two-sided *α* = 0.05 to detect absolute mean changes of 0.41 GAS-7 points (*d*_*z*_ = 0.15) in the Meliora group and 0.37 points (*d*_*z*_ = 0.15) in the Sham group. In a supportive analysis of the TAU group, GAS-7 scores decreased by 0.64 points from baseline to week 12 (95% CI, −0.92 to −0.35; *d*_*z*_ = −0.25; Holm-adjusted *p* = 4.2×10^−5^). Descriptive GAS-7 scores at baseline, post-intervention, and the week-24 follow-up for both the ITT and PP cohorts are presented in Table 1.

**Fig 1:**
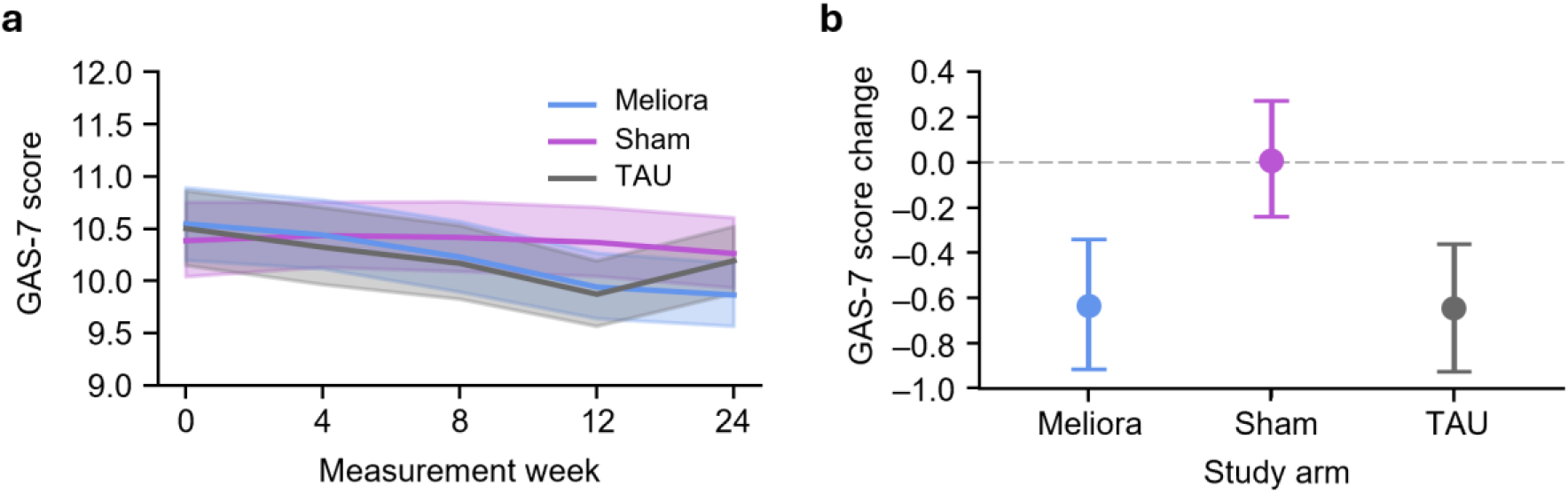
Game Addiction Scale (GAS-7) scores from baseline (week 0) to post-intervention (week 12) and follow-up (week 24) in the intention-to-treat (ITT) cohort. **a**. Mean GAS-7 scores over the 24-week study period across the three study arms. **b**. Change in GAS-7 scores from baseline to post-intervention (12 weeks). Shaded areas and error bars represent 95% bootstrapped confidence intervals.

**Table 1.** GAS-7 scores (mean, 95% bootstrap CI) for intention-to-treat (ITT) and per-protocol (PP) cohorts collected at baseline (week 0), post-intervention (week 12), and follow-up (week 24).

| Cohort | Arm | Baseline (week 0) | Post-intervention<br>(week 12) | Follow-up<br>(week 24) |
| --- | --- | --- | --- | --- |
| Intention-to-treat (ITT) | Meliora (n = 337) | 10.56 (10.22, 10.91) | 9.93 (9.49, 10.38) | 9.90 (9.43, 10.37) |
|  | Sham (n = 347) | 10.38 (10.04, 10.73) | 10.40 (10.07, 10.73) | 10.26 (9.94, 10.59) |
|  | TAU (n = 317) | 10.50 (10.15, 10.87) | 9.86 (9.41, 10.32) | 10.18 (9.73, 10.64) |
| Per-protocol (PP) | Meliora (n = 99) | 10.41 (9.83, 11.02) | 9.75 (9.24, 10.27) | 9.70 (9.12, 10.31) |
|  | Sham (n = 96) | 10.41 (9.76, 11.06) | 10.16 (9.55, 10.80) | 9.99 (9.40, 10.64) |
|  | TAU (n = 288) | 10.47 (10.10, 10.85) | 10.04 (9.69, 10.41) | 10.20 (9.87, 10.55) |

### Between-group comparisons

In supportive post hoc analyses using robust linear mixed modelling adjusted for baseline GAS-7 score, baseline PHQ-9 score, and demographic variables, GAS-7 change was more negative in the Meliora group than in the Sham group (adjusted mean difference = −0.19 points; 95% CI, −0.30 to −0.08; *d* = −0.13; Holm-adjusted *p* = 0.001), indicating a relatively greater GAS-7 reduction in the Meliora group than in the Sham group. GAS-7 change did not differ between the Meliora and TAU groups (adjusted mean difference = 0.04 points; 95% CI, −0.08 to 0.15; *d* = 0.03; Holm-adjusted *p* = 0.509). GAS-7 change was more positive in the Sham group than in the TAU group (adjusted mean difference = 0.23 points; 95% CI, 0.12 to 0.34; *d* = 0.16; Holm-adjusted *p* < 0.001), indicating a relatively greater reduction in the TAU group than in the Sham group. All tests were two-sided.

### Covariate analysis

From baseline to week 12, older age was associated with more positive GAS-7 change (adjusted mean difference = 0.09 points; 95% CI, 0.04 to 0.14; *d* = 0.06; FDR-adjusted *p* = 0.004), indicating relatively less reduction or greater increase in GAS-7 with increasing age. Higher income was associated with more negative GAS-7 change (adjusted mean difference = −0.10 points; 95% CI, −0.16 to −0.04; *d* = −0.07; FDR-adjusted *p* = 0.004), indicating relatively greater reduction or less increase with higher income. Higher baseline PHQ-9 scores were likewise associated with more negative GAS-7 change (adjusted mean difference = −0.02 points; 95% CI, −0.03 to −0.01; *d* = −0.01; FDR-adjusted *p* = 0.008). Effect sizes for all associations were small or negligible. No significant associations were observed for gender, education, or life status.

### Association between changes in depressive symptoms and GAS-7

To examine whether changes in gaming-related problems tracked depressive symptom changes, we examined associations between GAS-7 and PHQ-9 change from baseline to post-intervention. A small positive association was observed within the Meliora group (*r* = 0.17; *p* = 0.002), whereas associations were not statistically significant within the Sham group (*r* = 0.10; *p* = 0.062) or the TAU group (*r* = 0.07; *p* = 0.193). When all study arms were pooled, the association remained small but was statistically significant (*r* = 0.10; *p* = 0.001), with PHQ-9 change explaining 1.1% of the variance in GAS-7 change (*R*^*2*^ = 0.011).

### Intervention usage time and immersion

Intervention usage time was not significantly associated with change in GAS-7 scores from baseline to post-intervention in either the Meliora group (adjusted β per 1-SD higher use = −0.20; 95% CI, −0.45 to 0.06; d = −0.09; *p* = 0.125) or the Sham group (adjusted β = −0.11; 95% CI, −0.38 to 0.16; d = −0.05; *p* = 0.443). Similarly, experienced immersion, as measured with the Immersive Experiences Questionnaire, was not significantly associated with GAS-7 change in either the Meliora group (adjusted β per 1-SD higher immersion = 0.03; 95% CI, −0.23 to 0.30; d = 0.02; *p* = 0.804) or the Sham group (adjusted β = 0.15; 95% CI, −0.13 to 0.44; d = 0.07; *p* = 0.289). There was also no evidence of an interaction between intervention usage time and immersion in either the Meliora group (adjusted β = −0.13; 95% CI, −0.39 to 0.12; d = −0.06; *p* = 0.300) or the Sham group (adjusted β = −0.04; 95% CI, −0.28 to 0.21; d = −0.02; *p* = 0.772).

### Deterioration analysis

At the ≥25% threshold, deterioration from baseline to post-intervention occurred in 35/337 participants (10.4%) in the Meliora group, 53/347 participants (15.3%) in the Sham group, and 31/317 participants (9.8%) in the TAU group, with no statistically significant between-group differences (all pairwise Fisher exact tests, two-sided, Holm-adjusted *p* ≥ 0.107). Participants meeting this deterioration criterion had lower baseline GAS-7 scores than other participants in all three study arms (Meliora, 9.14 vs 10.72, FDR-adjusted *p* = 0.012; Sham, 8.87 vs 10.65, FDR-adjusted *p* < 0.001; TAU, 9.06 vs 10.65, FDR-adjusted *p* = 0.017), consistent with a relative-change criterion being more readily reached from lower baseline scores. Exploratory characterization analyses otherwise identified no consistent demographic or clinical profile across study arms. Lower baseline PHQ-9 scores were observed among deteriorators in the Sham group (13.91 vs 16.01, FDR-adjusted *p* = 0.017), and deteriorators in the TAU group were older than other participants (40.10 vs 32.66 years, FDR-adjusted *p* = 0.009), but neither association was observed consistently across study arms. Deterioration was not associated with male versus female gender in any study arm (all FDR-adjusted *p* ≥ 0.630). Neither intervention use nor experienced immersion differed between deteriorators and other participants in the Meliora or Sham group (all FDR-adjusted *p* ≥ 0.408). Consistently, greater intervention use was not significantly associated with deterioration in either the Meliora (OR per 1-SD higher use = 0.96; 95% CI, 0.66 to 1.40; *p* = 0.832) or the Sham group (OR = 1.04; 95% CI, 0.76 to 1.43; *p* = 0.802).

At the more stringent ≥50% threshold, deterioration occurred in 13/337 participants (3.9%) in the Meliora group, 21/347 participants (6.1%) in the Sham group, and 7/317 participants (2.2%) in the TAU group, again with no statistically significant between-group differences (all pairwise Fisher exact tests, two-sided, Holm-adjusted *p* ≥ 0.057). Given the small number of participants meeting this threshold, further subgroup characterization was not conducted because the resulting estimates would have been statistically unstable.

### Corroborative analyses in the per-protocol cohort

Finally, complementary analyses were conducted in the per-protocol cohort. Mean total intervention use was 45.5 h in the Meliora group and 44.9 h in the Sham group. The PP analyses were consistent with the ITT findings: despite substantially greater intervention exposure, mean GAS-7 scores did not increase in either the Meliora or Sham group, and no between-group differences suggesting greater worsening of GAS-7 scores in the Meliora group were observed (Supplementary Material 1).

## Discussion

Treatment dependency is a recognized concern in face-to-face treatments (20), but such problems have not typically been considered for DMHIs either with or without gaming elements (21,22). On the other hand, video games have been linked to gaming-related problems (23), which raises safety concerns for video game therapeutics. Here, ITT within-group comparisons showed a small decrease in GAS-7 scores from baseline to post-intervention in the Meliora group. While this score change was statistically significant, the effect size was small with a drop of less than 1 point on the GAS-7 scale indicating that the magnitude of the observed reduction was small. A similarly small decrease was observed in the TAU group, whereas no significant change was observed in the Sham group, despite similar intervention exposure in the Meliora and Sham groups. The deterioration analysis found no statistically significant between-group differences in individual-level worsening rates from baseline to post-intervention across Meliora, Sham, and TAU groups. This pattern of results was reproduced in the PP cohort, where a substantially greater intervention exposure was not accompanied by increases in mean GAS-7 scores. Together, these findings thus provide no evidence that long-term use of the video game therapeutic was associated with increased gaming-related problems in appropriately screened patients using a product with safeguards against excessive use.

Excessive entertainment gaming has been linked to gaming-related problems (24). Here, we found no association between intervention usage time and GAS-7 score changes. However, intervention exposure varied considerably, and average usage time in the ITT cohort was below the recommended 48 h over the 12-week intervention period. Although the per-protocol analyses examined participants with ≥24 h of intervention use, only 99 of 337 participants in the Meliora group and 96 of 347 participants in the Sham group met this criterion, limiting conclusions regarding consistently high intervention exposure. Importantly, participants with significant gaming- or gambling-related problems were excluded at baseline; therefore, these findings should not be generalized to patients with pre-existing problematic gaming or elevated susceptibility to gaming-related harms. Changes in entertainment gaming during the intervention period were not evaluated, and it remains unclear whether intervention use occurred in addition to or instead of entertainment gaming, and how intervention use may have influenced other gameplay and media use. Also, GAS-7 measured gaming-related problems related to digital games in general and did not specifically distinguish the studied interventions from other digital gaming. This broader assessment thus captured whether exposure to either intervention was associated with changes in participants’ overall gaming-related problems during the study period. Furthermore, no equivalence margin for GAS-7 was prespecified in the protocol for the present study. Therefore, these analyses cannot establish statistical equivalence or exclude the possibility of symptom increases smaller than those detectable with the available statistical power.

Gaming-related problems have also been associated with motivational and immersive aspects of gaming (25,26). In an earlier study of the present data, we found experienced immersion to be a key driver of therapeutic efficacy (10). However, experienced immersion was not associated with GAS-7 change in either intervention group, providing no evidence that greater experienced immersion was accompanied by increased gaming-related problems.

Previously, we assessed the safety of Meliora and Sham interventions using comprehensive open-ended self-report data on adverse events acquired with a novel multi-channel approach (17). An inductive analysis of these qualitative data revealed no reports of gaming-related problems (10), suggesting either an absence of subjectively experienced gaming-related problems, or unawareness of or underreporting of them. The present study provides complementary evidence using a self-report scale focused specifically on gaming-related problems. While these findings do not fully resolve this uncertainty, the structured GAS-7 assessments likewise did not indicate an increase in gaming-related problems during the study period.

In conclusion, we found no evidence that long-term use of a video game therapeutic increased gaming-related problems in appropriately screened patients and with interventions limiting excessive use. These findings provide converging albeit initial evidence that video game therapeutics can be used over extended periods without increasing gaming-related problems when safeguards for excessive use are incorporated into their design and implementation. Future studies should assess the risks of video game therapeutics in patients with prior gaming-related problems.

## Data Availability

Pseudonymized individual participant data underlying the results reported in this article will be considered for sharing upon reasonable request to the corresponding author following publication. Data sharing is subject to approval by the Helsinki University Hospital Regional Committee on Medical Research Ethics, compliance with the General Data Protection Regulation (GDPR) and other applicable laws, and execution of a data use agreement. Requests must include a scientifically and methodologically sound research proposal outlining the intended use of the data.

