## Supplementary material 1 for "No evidence of increased gaming-related problems with long-term use of a video game therapeutic: Exploratory endpoint findings from a randomized controlled trial"

### Per-protocol completer cohort analysis

A total of 483 participants were included in the per-protocol (PP) cohort (Meliora: $n = 99$; Sham: $n = 96$; TAU: $n = 288$). The criteria were to have filled at least the 8- or 12-weeks symptom assessment, and to have at least 24 h of intervention use within the intervention period. The mean age was 33.8 years (SD 9.5), and 66% were female. Baseline GAS-7 scores were similar between groups (Meliora: mean 10.41, 95% CI, 9.83 to 11.02; Sham: mean 10.41, 95% CI, 9.76 to 11.06; TAU: mean 10.47, 95% CI, 10.10 to 10.85). As reported earlier, in the per-protocol cohort, we observed no difference between Meliora and Sham groups in terms of intervention use (45.5 h, SD 16.0; 44.9 h, SD 17.6, respectively, $p = 0.80$), or subjective immersion (125.4, SD 25.0; 123.1, SD 24.7; $p = 0.53$).

Between baseline (week 0) and post-intervention (week 12), GAS-7 scores decreased by 0.67 points in the Meliora group (95% CI, −1.14 to −0.19; $d_z = -0.28$; Holm-adjusted $p = 0.012$) and by 0.43 points in the TAU group (95% CI, −0.72 to −0.14; $d_z = -0.17$; Holm-adjusted $p = 0.011$), whereas no statistically significant change was observed in the Sham group (mean change, −0.25 points; 95% CI, −0.73 to 0.23; $d_z = -0.11$; Holm-adjusted $p = 0.301$) (Table 1).

Using robust linear mixed modelling adjusted for baseline GAS-7 and PHQ-9 scores and demographic variables, no statistically significant differences in GAS-7 change were observed between Meliora and Sham groups (adjusted mean difference = −0.22 points; 95% CI, −0.50 to 0.06; $d = -0.18$; Holm-adjusted $p = 0.350$), Meliora and TAU groups (adjusted mean difference = −0.14 points; 95% CI, −0.36 to 0.09; $d = -0.11$; Holm-adjusted $p = 0.481$), or Sham and TAU groups (adjusted mean difference = 0.09 points; 95% CI, −0.14 to 0.32; $d = 0.07$; Holm-adjusted $p = 0.481$).

No associations with gender, baseline PHQ-9, age, education, income, or life status remained statistically significant after correction for multiple comparisons; effect sizes were small.

Using robust regression analysis adjusted for baseline GAS-7 and PHQ-9 scores and demographic variables, we found no significant associations between GAS-7 change from baseline to post-intervention and intervention use (Meliora: $p = 0.427$; Sham: $p = 0.914$) or experienced immersion (Meliora: $p = 0.607$; Sham: $p = 0.696$). There was also no evidence of an interaction between intervention use and immersion (Meliora: $p = 0.942$; Sham: $p = 0.237$).
